# Programming of the respiratory epithelium *in utero -* insights from the amniotic epithelial methylome

**DOI:** 10.1101/2025.10.02.25337047

**Authors:** Patricia Agudelo-Romero, Thomas Iosifidis, James Lim, Nina Kresoje, David G. Hancock, Guicheng Zhang, Abhinav Sharma, Talya Conradie, Minda Amin, Yuliya V. Karpievitch, Desiree T. Silva, Anthony Bosco, Susan L. Prescott, Peter N. LeSouëf, Elizabeth Kicic-Starcevich, Anthony Kicic, David J. Martino, Stephen M. Stick

## Abstract

**Background:** Dysregulation of the airway epithelium contributes to recurrent wheezing and asthma and may have developmental origins. Direct interrogation of fetal airway epithelium is not feasible in humans limiting investigation of prenatal epithelial programming.

**Methods:** We performed high-throughput target-capture DNA methylation sequencing of matched amniotic epithelial biopsies and neonatal nasal brushings from 84 mother-infant pairs within the Airway Epithelium Respiratory Illnesses and Allergy (AERIAL) cohort. We compared tissue-specific methylation landscapes for conservation and explored associations with gestational exposures known to influence childhood asthma risk (maternal asthma history and maternal prenatal smoking exposure).

**Results:** Between amniotic and nasal tissues, we identified 4,897 differentially methylated regions (FDR ≤ 0.05 and log_2_FC ≥ |0.2|) that were generally hypermethylated in the nasal epithelium. Despite these extensive tissue-specific differences, filtering for non-significant differential methylation loci (FDR ≥ 0.1) revealed 1,493,975 CpG loci (∼20%) with highly concordant methylation levels between tissues (Pearson’s R > 0.8). Within this conserved methylome fraction, we identified exposure-associated CpG sites linked to maternal asthma history (55 CpGs) and prenatal smoking exposure (164 CpGs) that showed consistent directionality in both tissues. These analyses were exploratory and limited by small numbers of exposed participants.

**Conclusions:** Amniotic epithelium shares a subset of conserved methylation features with the nasal epithelium, supporting its utility as a valuable surrogate for studying aspects of developmental programming of airway vulnerability. While exposure-associated signals were detectable within the conserved methylome fraction, these findings should be considered preliminary, but provide an encouraging framework for early risk stratification in asthma.

## Introduction

Asthma is a chronic inflammatory airway disease, among the most frequent causes of hospital admissions in children and represents a significant global disease burden (1–3). A substantial body of evidence suggests prenatal exposures play a critical role in shaping newborn lung function (4), and contribute to the airway epithelial cell dysfunction observed in children with asthma (5). Intrauterine exposure to air pollution and tobacco smoke are well-established risk factors for childhood asthma that impair fetal lung development and increase airway hyperresponsiveness in offspring (6). Similarly, poorly controlled maternal asthma in pregnancy has been associated with childhood wheeze and increased risk for asthma in offspring through alterations in fetal development (7). Animal models also suggest intrauterine chronic inflammatory conditions might directly influence fetal-placental development (8) and aberrant prenatal programming of the airway epithelium tissues (9). Understanding these developmental programming mechanisms is a research priority needed to facilitate early detection and targeted interventions.

Epigenetic factors such as DNA methylation play a crucial role in fetal lung development, and changes in DNA methylation have been observed in the airway epithelium in children with asthma (10, 11). Environmental exposures such as cigarette smoke and pollution induce reversible epigenetic changes in airway epithelial cells via oxidative stress (12, 13), disrupting developmental gene expression patterns, and contributing to asthma development (14, 15). Methylation may therefore be an important epigenetic mechanism through which to understand the complex interplay between the prenatal environment and fetal epithelial programming in the development of childhood asthma.

A key challenge to studying the prenatal origins of epithelial dysfunction relates to the challenges of sampling the newborn airway epithelial cells, which are difficult to obtain before key postnatal events occur, such as colonization of the mucosal surfaces, and exposure to airborne antigens. To address this, we explored the utility of fetal amniotic epithelial tissue as a surrogate for airway epithelial cells (16). The amnion is exposed to the maternal environment and may harbor epigenetic signatures (17, 18) that reflect processes occurring in the developing fetal respiratory tract. Amniotic epithelial cells possess stem cell like characteristics and are also attractive targets for cellular therapy (19). In this observational study, we explored the landscape of DNA methylation in amniotic epithelium and matched nasal epithelial samples from a cohort of healthy term newborns from the Airway Epithelium Respiratory Illnesses and Allergy (AERIAL) study (20). We conducted a comparative analysis of genome-wide DNA methylation profiles and examined the relative conservation of DNA methylation patterning at specific genes crucial for lung and epithelial development. We also explored whether methylation changes associations with key maternal exposures that influence asthma development were conserved in both tissues. Our results inform ongoing developmental programming studies on epithelium vulnerability.

## Methods

### Study design and samples

AERIAL (20) is a substudy nested within the ORIGINS birth cohort (21, 22). Matched newborn amnion biopsies, nasal epithelial brushings, and maternal urine were collected from 84 mother–infant pairs. Maternal asthma history and prenatal tobacco-smoke exposure were ascertained from at least two consecutive questionnaires administered at 20, 28, and 36 weeks’ gestation. The study was approved by the Ramsey Health Care HREC WA-SA (#1908) and written informed consent was obtained.

### Sample processing and DNA extraction

Placentas were processed within 48 hours after delivery (mean 20.4 ± 13.1 hours [SD]); with the chorion membrane manually separated (23) and amnion sampled. Newborn nasal brushings were collected within 6 weeks from birth (15.9 ± 7.9 days [SD]) (20). Samples were cryopreserved at −80°C. Genomic DNA was extracted from both tissues using the Chemagic 360 automated nucleic acid isolation system (Revvity, Baesweiler, Germany) and the Chemagic™ DNA Blood 400 Kit H96 (Revvity, part# CMG-1091), and quantified by fluorometry using the Qubit HS dsDNA Assay (Q32854, Thermo Fisher Scientific) (Figure E1 in the online supplement).

### Targeted DNA methylation sequencing

Targeted DNA methylation profiling was performed using enzymatic methyl-seq (EM-seq) with capture enrichment (TWIST Human Methylome Panel). Libraries were prepared from 200 ng genomic DNA, pooled pre-capture, enriched by hybridization capture, PCR amplified, and sequenced on an Illumina NovaSeq 6000 (paired-end 150 bp) (Figure E1 in the online supplement).

### Bioinformatics

FASTQ files were processed using nf-core/methylseq v23.04.2 (24) and aligned to GRCh38 with BWA-meth; methylation was called with MethylDackel and QC metrics generated using target-methylseq-qc v2.1.0 (25). Panel-specific interval filtering was applied prior to downstream analyses (Supplemental Methods; Figure E1). Analyses used ABLeS (26) and Pawsey resources (27, 28). Scripts are available at https://github.com/wal-yan/AERIAL/tree/main/RespEpithelium_Methylome.

### Statistical analysis

Analyses were performed in R v4.3.2 (RStudio v2023.03.0+386) (29) (Supplemental Methods; Figure E1). CpGs with zero coverage in any sample, low coverage (<5), or extreme coverage (>500) were removed; mitochondrial and non-standard chromosomes were excluded. Methylation was expressed as β = methylated/(methylated+unmethylated) ×100 and transformed to M values for statistical modelling. Sample QC included sex inference and principal component analysis. Tissue differences were assessed at the region (DMR) and CpG (DMP) levels using Benjamini–Hochberg FDR correction. DMRs were identified using DMRcate v2.16.1 (30) (tissue as primary predictor, adjusted for sex), requiring ≥50 CpGs per region (FDR ≤0.01; |mean difference| ≥1). DMPs were tested using limma v3.58 (31) with sex as a fixed effect and paired samples accounted for using *duplicateCorrelation()* with participant ID (FDR ≤0.05; |log2FC| ≥0.2). Conserved CpGs were defined as non-differential between tissues (FDR ≥0.1) with high concordance (Pearson R ≥0.8). Maternal asthma and in utero smoke exposures were tested at conserved CpGs using limma (adjusted for sex; participant ID block; FDR ≤0.05; |log2FC| ≥0.2). CpGs/regions were annotated to genes, and GO enrichment was assessed using GREAT-based analyses v2.4.0 (32). Cell deconvolution was not performed as validated reference methylomes for amniotic epithelium are not currently available and inclusion of surrogate variables introduced instability in the paired modelling framework.

### Urinary cotinine

Maternal smoking was self-reported and validated using urinary cotinine (33) measured at 20 and 36 weeks’ gestation (21, 22) with the Salimetrics® High Sensitivity Salivary Cotinine Enzyme Immunoassay Kit (Cat No. 1-2002-5; Salimetrics, State College, PA, USA). Cotinine >10 ng/mL indicated active smoking.

## Results

### Demographics and Sequencing quality

The characteristics of the study population are presented in Table 1. While the newborns’ ethnic backgrounds were diverse, the cohort was predominantly Caucasian, with 51.2% of mothers and 48.8% of fathers identifying as such. Newborns had a mean gestational age of 39.12 ± 1.28 weeks [SD], with three preterm births recorded in the cohort, occurring between 35 and 36 weeks of gestation. Sequencing quality statistics were similar across tissues (Table 2), with low duplication rates and 93% of bases covered at a depth of at least 30X. The average number of paired-end sequences generated was approximately 183 ± 28.90 [SD] million for amnion samples and 168.67 ± 33.50 [SD] million for nasal samples. The methylation status was assessed across 7,936,546 million individual CpG dinucleotides, with 6,961,516 remaining after quality control filtering. Fold-80 base penalty metrics were between 1.2-1.6 across all samples indicating good uniformity of coverage for low-diversity libraries.

**Table 1.**
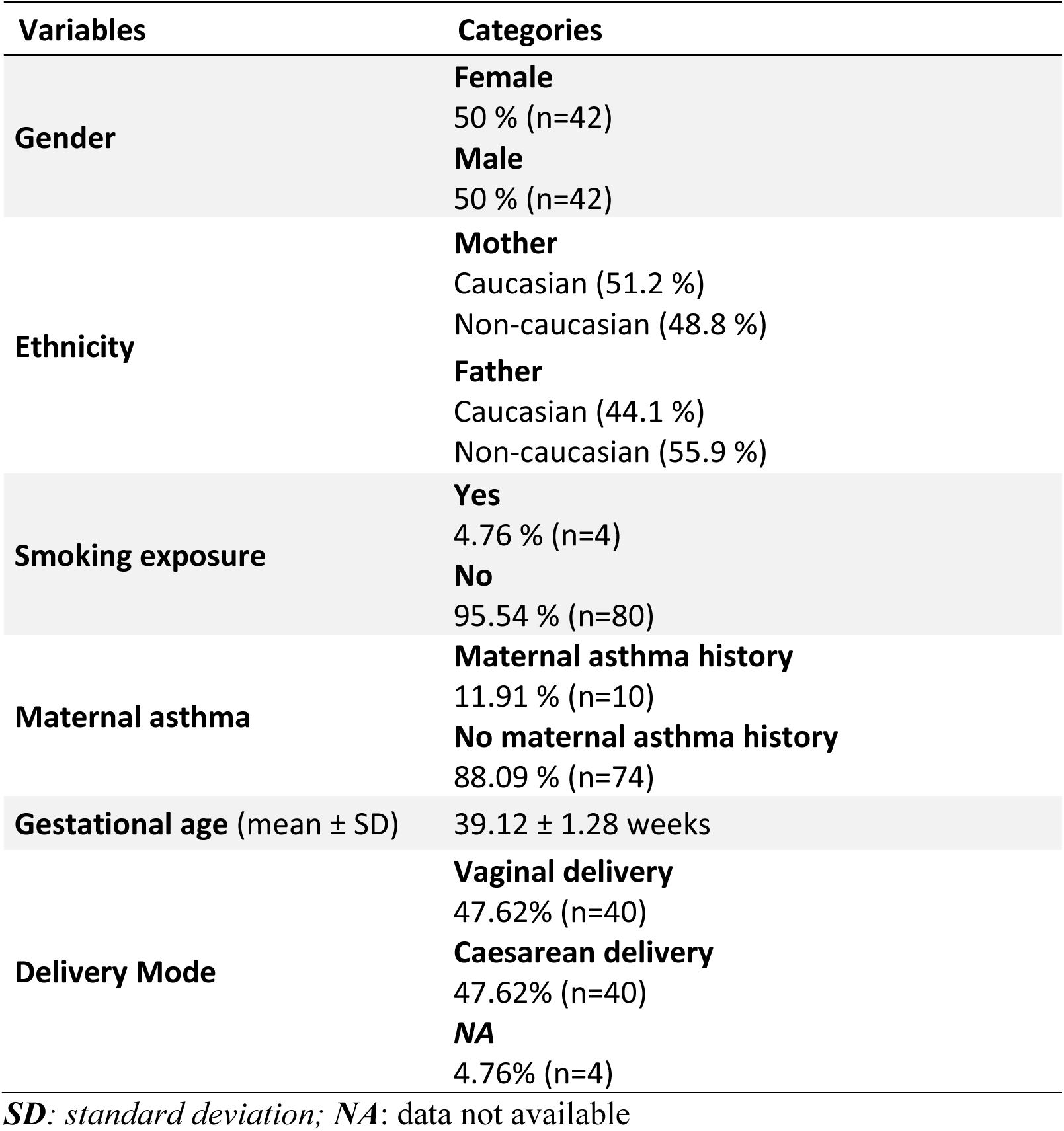
Demographic characteristics of the Airway Epithelium Respiratory Illnesses and Allergy (AERIAL) study.

**Table 2.** Sequencing statistics for 84 matched samples from neonatal nasal and maternal amnion. Values represent the mean ± standard deviation.

|  | Nasal samples | Amnion samples |
| --- | --- | --- |
| **M seq** (in million) | 168.67 ± 33.50 | 183 ± 28.90 |
| **% GC** | 27 ± 0.01 | 27% ± 0.01 |
| **% Duplication** | 15% ± 0.03 | 18% ± 0.03 |
| **Insert Size** | 225.35 bp ± 21.02 | 212.95 bp ± 9.98 |
| **% Target Bases 10X** | 99% ± 0.00 | 99% ± 0.00 |
| **% Target Bases 30X** | 93% ± 5.73 | 93% ± 8.55 |

bp: base pair

### Differential Methylation Landscape Between Amniotic and Nasal Tissues

We began by examining the general features of epithelium methylation landscapes between amniotic and nasal tissues, which revealed similar enrichment of methylated CpG sites around transcriptional start sites (Figure 1A), and the distribution of methylated and unmethylated CpG islands was similar across tissues (Figure 1B). These patterns were consistent across gene promoters and other regulatory features such as gene enhancers (Figure 1A and Figure E2A in the online supplement). Despite the overall similarities in CpG enrichment, dimensional reduction analysis of all 6,961,516 CpG sites across the genome revealed tissue type as the primary source of variation in DNA methylation profiles (PERMANOVA: pseudo-F=91.52, R^2^ 0.36, p-value=0.001) suggesting substantial tissue-specific differences (Figure 1C).

**Figure 1.**
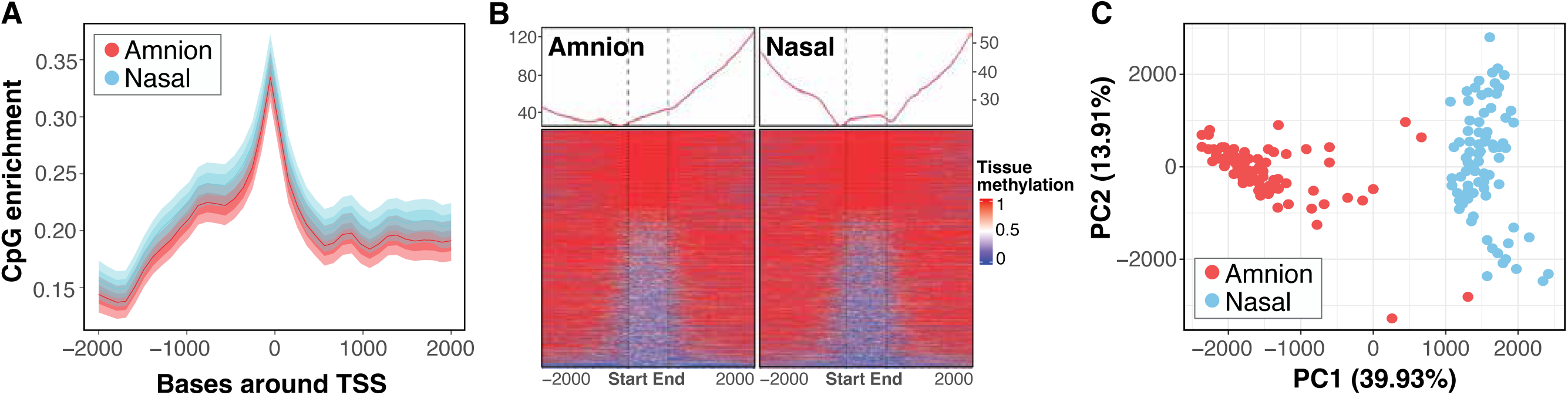
Comparative analysis of DNA methylation landscapes in amnion and nasal tissues. (A) Density plot illustrating CpG enrichment around Transcriptional Start Sites (TSSs). (B) Methylation levels of CpG islands with ± 2 kb. CpG islands are rescaled to the same width from Start to End vertical lines. (C) Principal component analysis (PCA) of genome-wide DNA methylation tissue profiles reveals distinct clustering of Amnion (red) and Nasal (blue) samples. Principal Component 1 (PC1) accounts for 39.93% of the variance.

To determine the extent of tissue-specific differences in DNA methylation patterns, we conducted differential analysis comparing amniotic and nasal epithelium at individual CpG sites and identifying the differentially methylated regions (DMR). This analysis revealed 716,075 individual CpGs that contributed to 4,897 genome-wide significant DMRs (Table E1 in the online supplement). Notably, in the nasal epithelium most regions exhibited higher methylation, at 3,847 regions (78.50%), while 1,050 regions (21.50%) showed a loss in methylation, compared to the amniotic epithelium. The volcano plot highlights the most hypomethylated region in amnion relative to nasal tissue as an intergenic region on chromosome 15 (chr15:70473876-70475721), while the most hypermethylated region was within the *NAXD* gene on chromosome 13 (chr13:110626965-110629183) (Figure 2A). Genomic track visualization revealed broad and consistent methylation differences across these loci (Figure 2B). The intergenic region on chromosome 15 exhibited higher methylation levels in amnion, whereas *NAXD* on chromosome 13 showed increased methylation in nasal tissue (Figure 2B). To gain broader insights into the biological processes associated with these methylation differences, we annotated DMRs to the nearest genes and performed Gene Ontology (GO) enrichment analysis. This revealed strong enrichment of pathways related to development and differentiation, with additional ontologies related to stimuli and metabolic processes (Figure 2C and Figure E2B in the online supplement).

**Figure 2.**
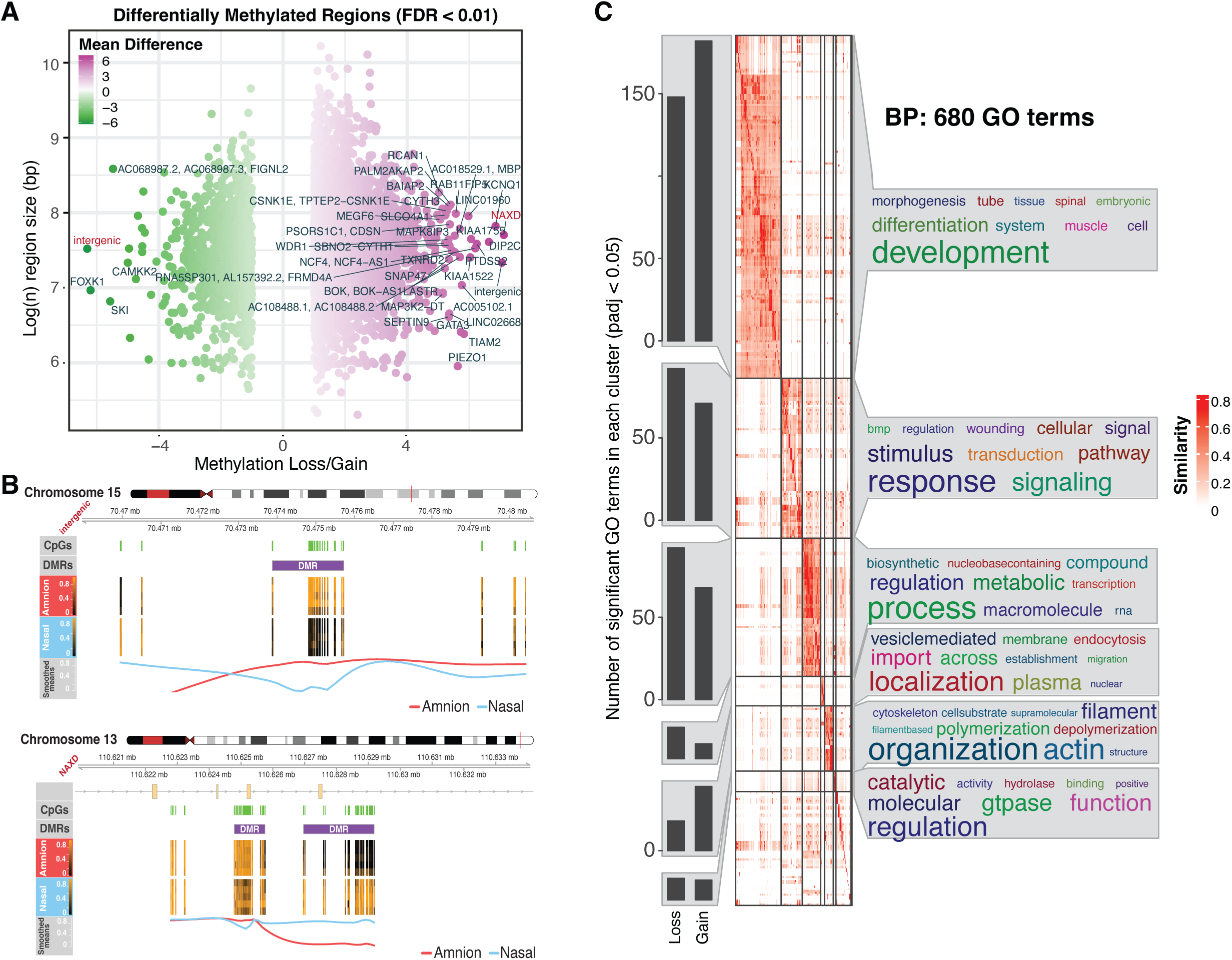
Regions differentially methylated between amniotic and nasal samples are enriched in cell development pathways. (A) Volcano plot displaying differentially methylated regions (DMRs) between amnion and nasal tissues. The x-axis represents the mean methylation difference, and the y-axis represents the log_2_-transformed region size. Genes associated with the most significant DMRs are labelled in red. (B) Genomic tracks showing methylation loss or gain in specific regions of Chromosomes 13 (*NAXD*) and 15 (Intergenic region), respectively. CpG islands are marked in green. The line graph summarizes the methylation difference between amnion (red) and nasal (blue) tissues across the region. (C) Gene Ontology (GO) enrichment was calculated using genes near DMRs between amniotic and nasal tissues, followed by a similarity matrix analysis. The similarity heatmap clusters significant GO terms into related Biological Process (BP), with colour intensity representing the significance of enrichment. Similarity of related terms within the same cluster is shown in the heatmap.

### Conservation of Methylation Across A Subset of Lung Specific Genes

We identified a conserved methylation signature (Figure 3A and Table E2 in the online supplement), comprising 1,493,975 CpG loci, representing approximately 21% of the total CpG landscape (n=6,961,516). This conserved pattern was distributed across all genomic regulatory features, with the highest representation in introns (20.09%), CpG islands (14.10%), and promoters (12.10%) (Figure E2A in the online supplement). To explore whether there are conserved methylation patterns in genes biologically relevant to epithelial and lung development, we queried the Genotype-Tissue Expression (GTEx) consortium database (https://gtexportal.org/home/) to extract a set of genes uniquely expressed in lung tissue compared to all other anatomical sites (total 459 genes). Among these gene sets, 159 genes (∼34.6%) exhibited highly concordant methylation patterns (Pearson’s R ≥ 0.8) between the two tissues and general conservation of the methylation landscape (Figure 3B). For instance, Surfactant Protein A1 (*SFTPA1)* exhibited intermediate levels of DNA methylation in both amnion and nasal tissues (Figure E2B in the online supplement), while the epithelial specific gene, Secretoglobin Family 3A Member 1 (*SCGB3A1),* known to regulate cell proliferation was predominantly hypomethylated in both tissues (Figure E2B in the online supplement), indicating conservation in subsets of biologically relevant genes. We conducted a pathway enrichment analysis of the conserved signature revealing enrichment of terms related to anatomical and morphogenic processes, cellular organization, metabolism and cell cycling, as well as the regulation of signal transduction and stimulus response (Figure 3D and Figure E2C in the online supplement).

**Figure 3.**
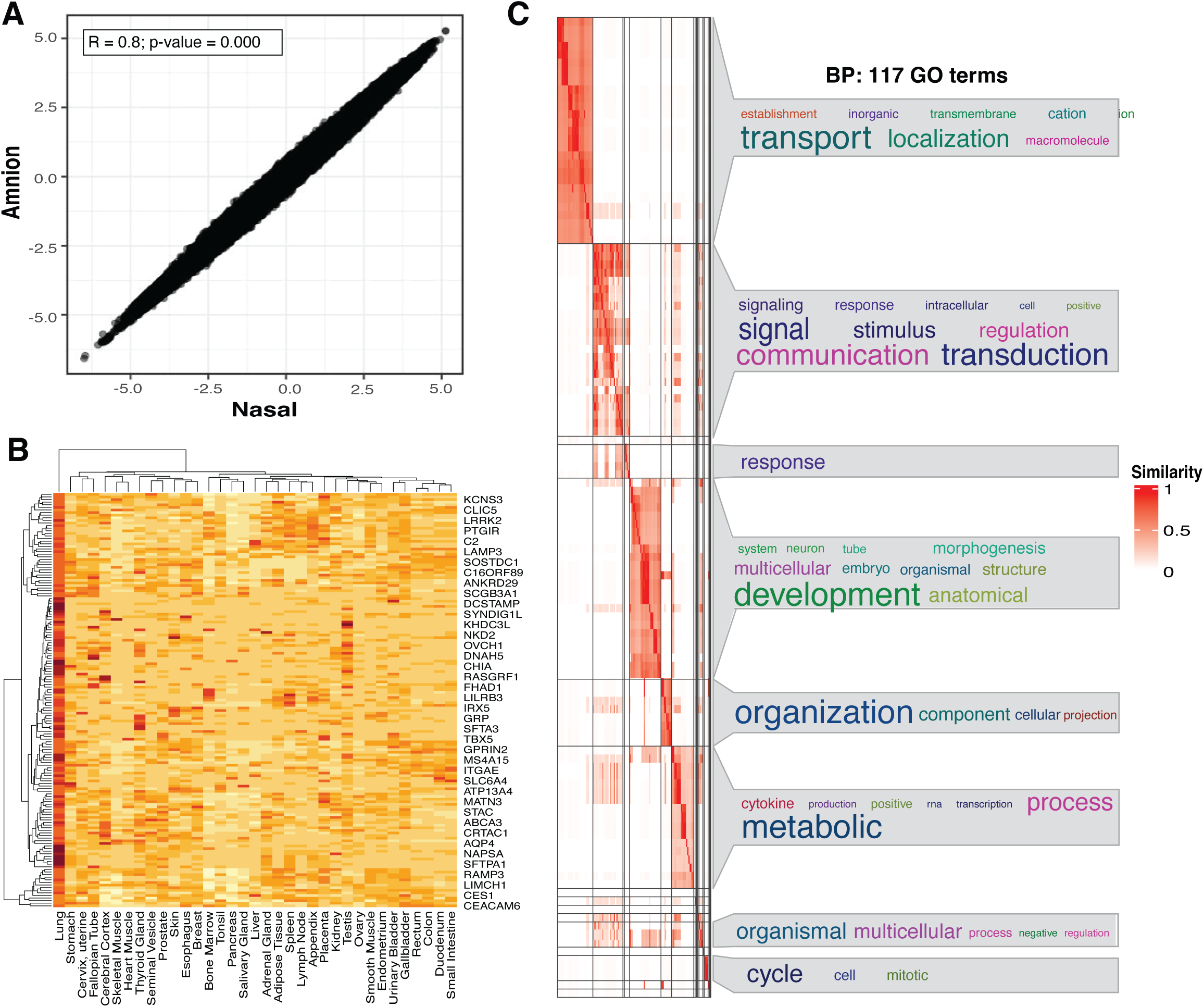
Conserved methylation landscapes were enriched in cell differentiation and morphogenesis pathways. (A) Scatter plot comparing DNA methylation levels between amnion and nasal tissues. Each dot represents a genomic region, with its position indicating the methylation level in each tissue. The strong positive correlation (Pearson’s R ≥ 0.8, p-value = 0.000) suggests a high degree of conservation in methylation patterns between these tissues. (B) Heatmap illustrating the expression levels of lung- and epithelial-specific genes across various tissues. Each row represents a gene, and each column represents a tissue. The colour intensity indicates the level of expression, with red representing high expression and blue representing low expression. Data are derived from the GTEx catalogue. (C) Gene Ontology (GO) enrichment was calculated using genes near CpG sites from the conserved methylation signature, followed by a similarity matrix analysis. The similarity heatmap displays GO terms associated with Biological Process (BP), with colour intensity representing the significance of enrichment.

### Amnion tissue preserves methylation signatures associated with exposures during pregnancy

Within the conserved portion of the methylome (Table E2 in the online supplement), we investigated statistical associations with two well-characterized exposures known to increase the risk of asthma development in offspring: maternal asthma history and *in utero s*moking exposure (34). In this cohort, 9 out of 84 mothers reported a history of asthma. Binary logistic regression analysis, adjusted for multiple comparisons (FDR ≤ 0.05), identified a small but significant associations at 55 CpG sites (51 hypermethylated and 4 hypomethylated) with maternal asthma (Table 3 and Table E3 in the online supplement). Principal component analysis (PCA) of the conserved CpG sites was significantly associated with maternal asthma history and this exposure was the major driver of variation in DNA methylation at these CpGs in both tissues (PERMANOVA: pseudo-F=27.03, R^2^ 0.14, p-value=0.0001) (Figure 4A). Enrichment analysis linked these DMPs to pathways involved in cellular response to stimulus, organization and development (Figure E3 in the online supplement). We applied Partial Least Squares - Discriminant Analysis (PLS-DA) for assessing the most relevant features, which identified the 15 CpGs contributing to group separation including four CpGs annotated to *MTHFSD* (Figure 4B). Consistent with the separation, the most discriminative CpGs showed lower methylation (M-values) in the maternal asthma group, with similar patterns across both tissues. We visualized the top three CpG sites shown in Figure 4C, chr16:86536881 (*MTHFSD*), chr16:86537583 (*MTHFSD*), and chr12:22507624 (*C2CD5/C2CD5-AS1*), which exhibited consistent differences in both amnion and nasal tissues in relation to maternal asthma history.

**Figure 4.**
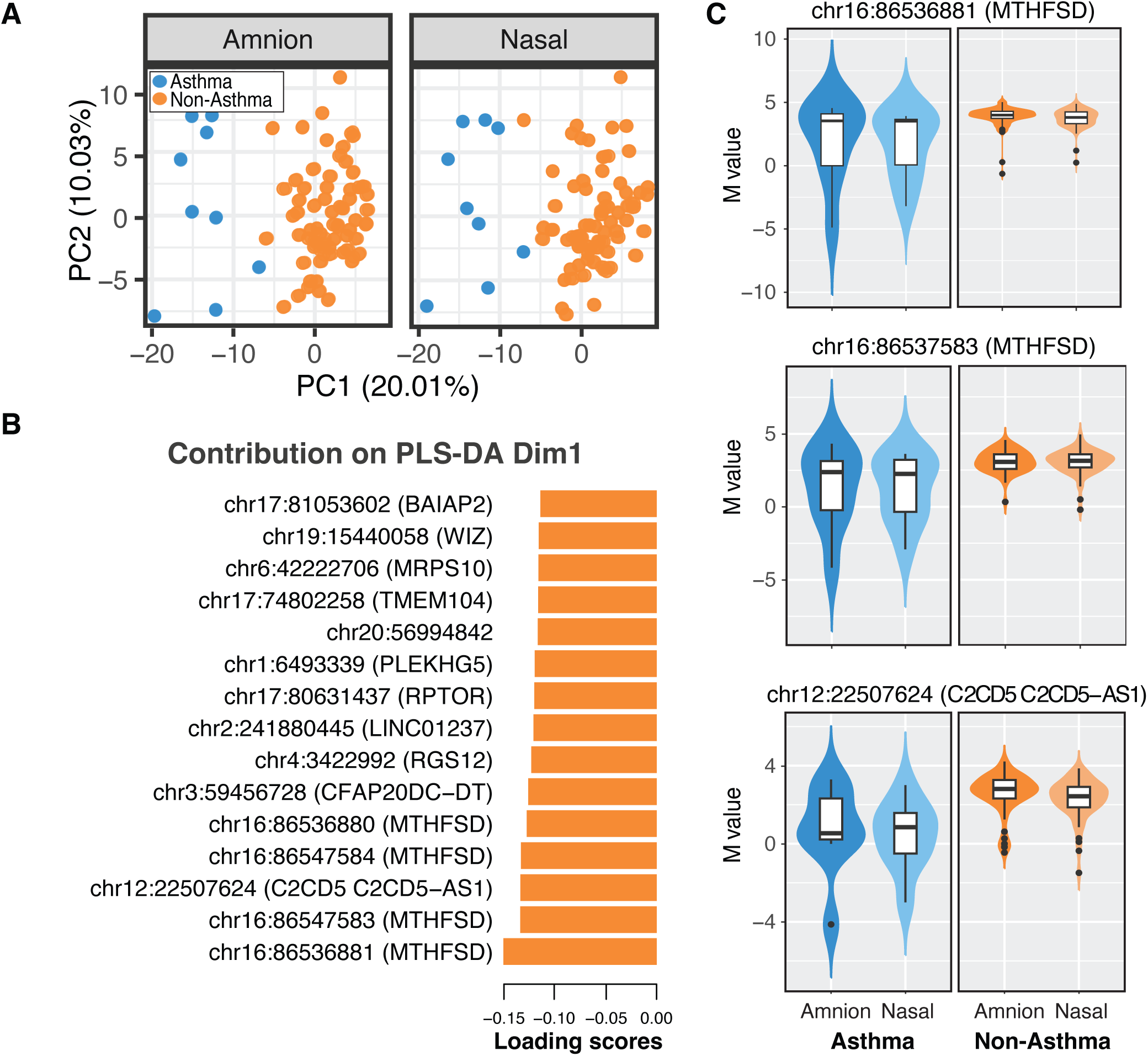
Methylation changes associated with maternal asthma in the conserved signature. (A) Principal Component Analysis (PCA) of DNA methylation variation in newborn amnion and nasal tissues, showing the first two principal components (PC1 and PC2). Samples are coloured based on maternal asthma history, with asthma in orange and non-asthma in blue. (B) Bar plot of loading scores from Dimension 1 (Dim1) of a Partial Least Squares Discriminant Analysis (PLS-DA), highlighting the top 15 differentially methylated CpG sites associated with maternal asthma. (C) Violin plots of the top three CpG sites identified by PLS-DA, showing the distribution of methylation levels (expressed as M-values) at features associated with maternal asthma in newborn amnion and nasal tissues, further stratified by maternal asthma history during pregnancy status.

**Table 3.** Top 10 Most Differentially Methylated Positions (DMPs) in Maternal Asthma History.

| Position | Symbol* | Log<sub>2</sub>FC | Adj. p-value |
| --- | --- | --- | --- |
| chr16:89629808 | *DPEP1* | 3.36 | 0.03172786 |
| chr16:89629809 | *DPEP1* | 3.02 | 0.02803976 |
| chr12:109788572 | *TRPV4* | 2.99 | 0.02178272 |
| chr5:72381577 |  | 2.88 | 0.02803976 |
| chr5:72381576 |  | 2.85 | 0.03384522 |
| chr12:109777366 | *FAM222A-AS1* | 2.85 | 0.008949334 |
| chr1:25812646 | *MTFR1L*; *SELENON* | -2.43 | 0.02803976 |
| chr1:25813427 | *MTFR1L*; *SELENON* | -2.47 | 0.03615935 |
| chr6:166412533 | *RPS6KA2* | -2.92 | 0.04016408 |
| chr6:166412532 | *RPS6KA2* | -3.36 | 0.03419868 |

\*CpG sites without a gene symbol may be located in regions with incomplete or less characterized gene annotations.

Next, we used the urinary cotinine measurements at 20 and 36 weeks of gestation as a marker of intrauterine smoke exposure. In our cohort, 4 of 88 mothers had cotinine levels consistent with tobacco smoking. Binary logistic regression analysis identified 164 CpG significant associations (126 hypermethylated and 38 hypomethylated) with maternal smoking exposure (Table 4 and Table E4 in the online supplement). Principal component analysis (PCA) of the conserved CpG sites significantly associated with smoking exposure identified it as primary source of the variation in DNA methylation in both tissues (PERMANOVA: pseudo-F=27.23, R^2^ 0.15, p-value=0.001) (Figure 5A), with significant enrichment of pathways involved in regulation of localization, cell activation, proliferation, and lung development (Figure E4 in the online supplement). By applying PLS-DA the 15 highly ranked features contributing to group separation (Figure 5B) comprised 14 CpG sites with decreased levels in the exposed group, while one CpG increased. Visualization of the top three significant CpG sites: chr12:116575870 (*MAP1LC3B2*), chr14:74362734, and chr17:36093007 (*CCL3*), also demonstrated the consistency of these changes across both amnion and nasal tissues between individuals with and without prenatal smoking exposure (Figure 5C).

**Figure 5.**
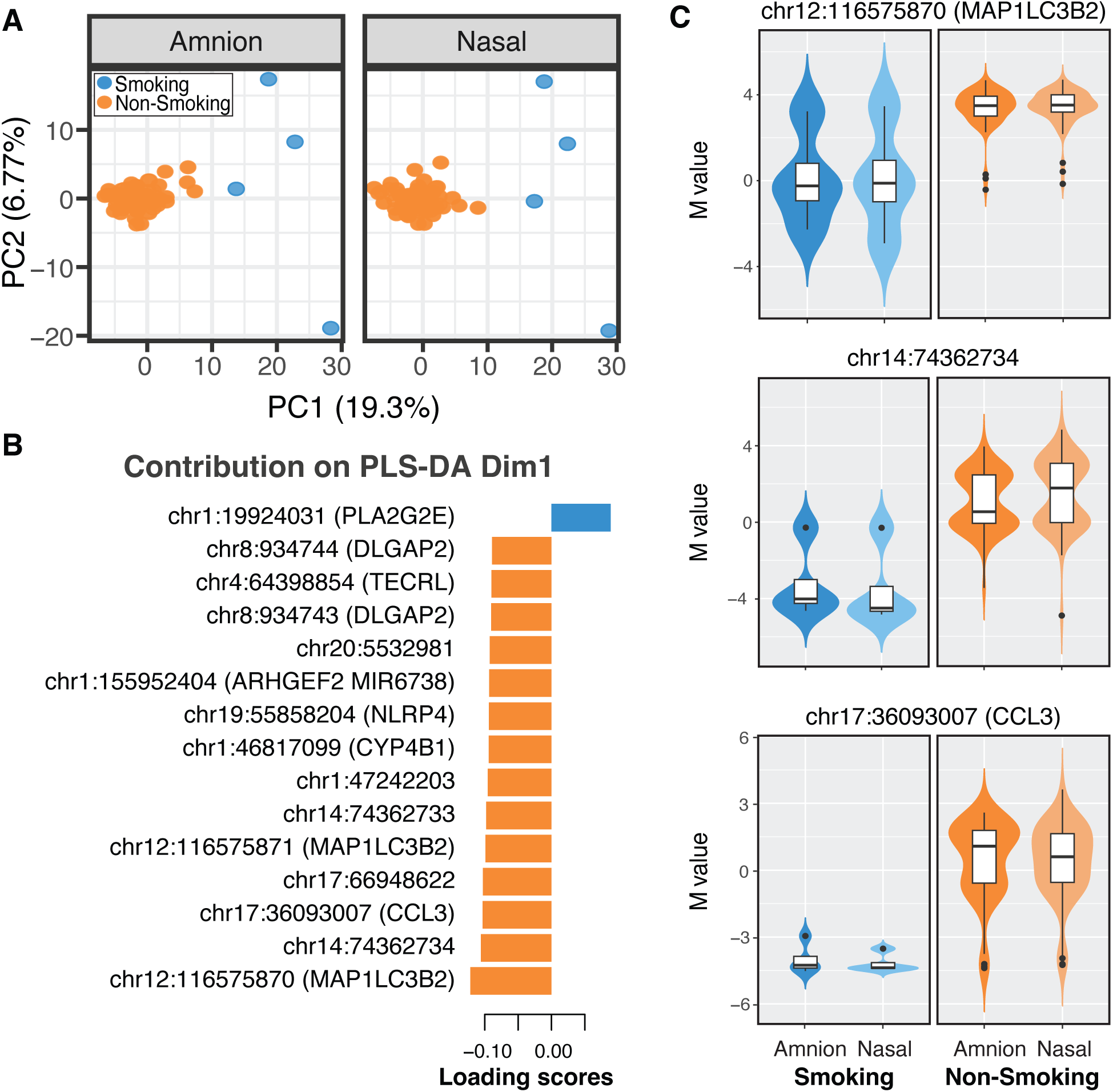
Methylation changes associated with maternal smoking exposure in the conserved signature. (A) Principal Component Analysis (PCA) of DNA methylation variation in newborn amnion and nasal tissues, showing the first two principal components (PC1 and PC2). Samples are coloured based on maternal smoking exposure, with smoking exposure in orange and non-smoking exposure in blue. (B) Bar plot of loading scores from Dimension 1 (Dim1) of a Partial Least Squares Discriminant Analysis (PLS-DA), highlighting the top 15 differentially methylated CpG sites associated with maternal smoking exposure. (C) Violin plots of the top three CpG sites identified by PLS-DA, showing the distribution of methylation levels (expressed as M-values) at features associated with maternal smoking exposure in newborn amnion and nasal tissues, further stratified by prenatal smoking status.

**Table 4.**
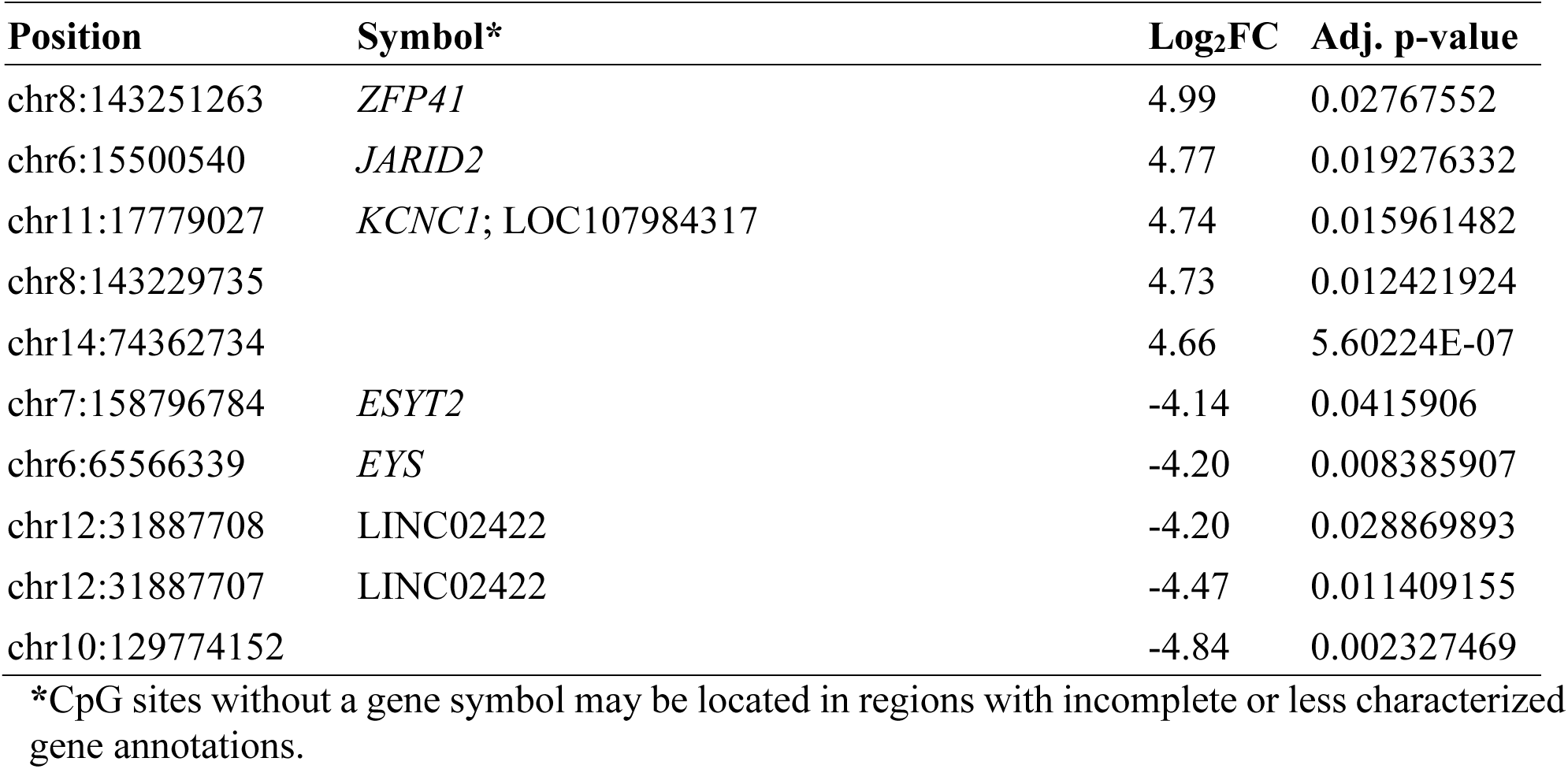
Top 10 Most Differentially Methylated Positions (DMPs) in Maternal Smoking Exposure.

## Discussion

In this study we evaluated the extent to which amniotic epithelium reflects DNA methylation features of the neonatal nasal airway epithelium, with the goal of assessing its utility for studying fetal programming of epithelial function relevant asthma development. To achieve this, we investigated the relationship between the amniotic and nasal epithelial methylation landscapes in ex vivo samples from newborns from a well characterized cohort. Our data suggest a sizeable fraction of CpG loci shows highly concordant methylation levels across tissues, enriched for genes involved in developmentally relevant pathways suggesting shared regulatory features established in fetal development.

Substantial tissue-specific methylation differences existed between the two tissues, characterized by a predominantly hypomethylated profile (reduced methylation) in amniotic epithelium samples. Given that DNA methylation levels typically increase in more terminally differentiated cell types, reflecting a more restrictive chromatin structure, this hypomethylation in amniotic tissue aligns with its more pluripotent state and stem-cell like characteristics reported in regenerative medicine studies (19). Despite these considerable differences, general features of the methylation landscape were similar, including enrichment of methylated CpGs around transcriptional start sites and low methylation across CpG Islands. consistent with patterns reported across multiple human somatic tissues (35). At the same time, histone modifications have been reported to contribute to epithelial plasticity, which is involved in both development and disease progression (36, 37). This divergence in epigenetic control may facilitate tissue-specific adaptations and responses to environmental changes, underscoring the dynamic nature of the epigenome.

Around 20% of the regions captured by the EM-seq target capture assay exhibited very high concordance of methylation levels, which we deemed the ‘conserved methylome’ between amniotic and nasal tissues (Figure 3A, Table E2 in the online supplement). This shared methylation signature may reflect common developmental and regulatory pathways shaped by the established during fetal development. Importantly these conserved loci imply a restricted subset of the methylome that may capture shared developmental or regulatory programs. Amniotic fluid contains bioactive molecules that play essential roles in fetal development, particularly by influencing critical cellular processes in lung epithelial cells (38). Additionally, the amniotic membrane contributes to the production of surfactant proteins and lipids vital for fetal lung maturation (39). Notably, within this conserved fraction, genes that play a critical role in lung development were represented, supporting the utility for amniotic tissue as a window for developmental programming of the respiratory epithelium (Figure 3B). However, the extent to which additional biologically relevant genes and loci fall outside the regions interrogated remains unknown. Additionally, the temporal separation between tissue sampling (up to six-week post-delivery for nasal samples) which reflects practical challenges in cohort studies means we cannot exclude the possibility that some methylation features in the nasal epithelium reflect early postnatal exposures. However this would be expected to reduce cross-tissue concordance, and the stability of the conserved CpG identified may reflect marks established prenatally that persist into the early postnatal period. Future studies using more comprehensive and unbiased methods, such as whole-genome bisulfite sequencing, could help uncover additional loci of interest.

Within the conserved methylome fraction, we conduced exploratory analyses to assess whether selected maternal exposures known to influence the development of asthma were reflected in both tissues (Figure 5 and Figure 6). Associations with maternal asthma history and prenatal smoking exposure were identified at a limited number of CpG loci, with directionally consistent effects observed in amniotic and nasal epithelium (Table E3 in the online supplement). These findings suggest that some exposure-related epigenetic signals established in utero may persist within shared epithelial regulatory regions after birth, consistent with previous studies (40, 41).

These exposure analyses must be interpreted cautiously. The number of exposed participants was small limiting statistical power and increasing susceptibility to outlier effects. Our exposure testing was intentionally restricted to the conserved methylome fraction, precluding inference regarding tissue-specific exposure effects or overall concordance across the epithelium. We leveraged PLS-DA purely for data visualization and descriptive summaries, not for feature selection or classification. We also acknowledge the observational nature of this study precludes establishing causality. These results are therefore best viewed as hypothesis-generating signals that motivate future studies in larger cohorts.

In conclusion, this study demonstrates that amniotic epithelium shares a restricted but biologically meaningful subset of DNA methylation features with the neonatal nasal airway epithelium. These support the use of amniotic tissue as a complimentary tissue to assess aspects of prenatal programming, providing opportunity for early risk stratification when direct sampling of airway epithelial cells from newborns is not feasible. Larger longitudinal studies incorporating clinical respiratory outcomes are planned to further establish the relevance of exposure-associated methylation signals for asthma risk stratification.

## Data availability

Raw sequencing data cannot be shared due to ethical/privacy restrictions. De-identified methylation β and M values for participants who provided genomics data sharing consent are deposited in Mendeley Data (https://data.mendeley.com/datasets/xsch4ysfsn/1). De-identified data for these participants are available from the authors upon reasonable request.

## Acknowledgments

We would like to thank the contributions of the AERIAL families, together with the cohort study investigator Jose A. Caparros-Martin, Lidija Turkovic, and our consumers Rael Rivers, Kate McGee and Olivia Gleeson, and the Western Australian Epithelial Research Program Consumer Reference Group for their contributions towards project inception/design and its adaptation through COVID-19 and other related amendments.

We also extend our gratitude to the hardworking and dedicated AERIAL research team for recruitment, liaising, and sample collection and processing over the duration of this study, with special mention to Bailee Renouf, Courtney Kidd and Ashleigh Heng-Chin.

We are grateful to all the ORIGINS families who support the project. We would also like to acknowledge and thank the following teams and individuals who have made ORIGINS possible: ORIGINS project team; Joondalup Health Campus (JHC); members of ORIGINS Community Reference and Participant Reference Groups; Research Interest Groups and the ORIGINS Scientific Committee; The Kids Research Institute Australia; City of Wanneroo; City of Joondalup; and Professor Fiona Stanley.

This study is a sub-project of ORIGINS. This unique long-term study, a collaboration between The Kids Research Institute Australia and Joondalup Health Campus, is one of the most comprehensive studies of pregnant women and their families in Australia to date, recruiting 10,000 families over a decade from the Joondalup and Wanneroo communities of Western Australia.

