## Supplemental Material for "Programming of the respiratory epithelium *in utero -* insights from the amniotic epithelial methylome"

### ***Supplementary Material***

### Supplemental Methods.

**Figure E1. Analysis Workflow.** This study was conducted in three main phases: Sample processing and sequencing (green), Bioinformatics analysis (blue), and Statistical analysis (pink). The statistical analysis phase was further divided into four key steps: Step 1 – Dimensional reduction using Principal Component Analysis (PCA); Step 2 – Differential methylation analysis at the regional level (differentially methylated regions, DMRs) associated with tissue differences; Step 3 – Differential methylation analysis at the CpG site level (differentially methylated positions, DMPs) associated with tissue differences; Step 4 – Identification of a conserved DMP signature, which was then used to assess associations with gestational exposures (maternal asthma and maternal smoking). MD: Mean Difference; FDR: False Discovery Rate; FC: Fold Change.

**Figure E2. Conserved methylation landscape across tissues. (**A) Histogram counts of CpGs annotated to genomic features for the conserved methylome. (B) Significantly enriched biological processes within the conserved methylome for CpGs annotate to nearest transcriptional start sites. Point size represents count of number of genes relative to total genes within the pathway (C) Raw sequencing coverage across the SFTPA1 genet coding region (top panel) and the SCGB3A1 coding region. Ticks in red are methylated reads and blue represents unmethylated reads. Height of the grey silhouettes represents sequencing coverage.

**Figure E3. Pathway enrichment of maternal asthma history.** (A) Significantly enriched biological processes within the conserved methylome for CpGs associated with maternal asthma. Point size represents count of number of genes relative to total genes within the pathway. (B) Gene Ontology (GO) enrichment was calculated using genes near CpG sites from the maternal asthma signature, followed by a similarity matrix analysis. The similarity heatmap displays GO terms associated with Biological Process (BP), with colour intensity representing the significance of enrichment.

**Figure E4. Pathway enrichment of maternal smoking exposure.** (A) Significantly enriched biological processes within the conserved methylome for CpGs associated with maternal smoking. Point size represents count of number of genes relative to total genes within the pathway. (B) Gene Ontology (GO) enrichment was calculated using genes near CpG sites from the maternal smoking signature, followed by a similarity matrix analysis. The similarity heatmap displays GO terms associated with Biological Process (BP), with colour intensity representing the significance of enrichment.

### Supplemental Methods

#### Study Design

AERIAL (1) is a prospective birth cohort nested within The ORIGINS birth cohort (2, 3). Matched pairs of amniotic membrane biopsies, nasal brushings and maternal urine were obtained from 84 newborns and their mothers. Maternal asthma history and tobacco smoke exposure during pregnancy was ascertained from at least 2 consecutive questionnaires administered at 20, 28 and 36 weeks of pregnancy. This study was conducted in accordance with the Declaration of Helsinki and was approved by the Ramsey Health Care HREC WA-SA (#1908). All parents, guardians, or next of kin provided written informed consent to participate in this study and collect maternal and newborn samples for the downstream analyses presented in this article.

#### Sample Collection and Processing

Placentas were processed within 48 hours post-birth on average (20.4 ± 13.1 hours [SD (Standard Deviation)]), with the chorion membrane manually separated (4) and the amnion membrane sampled. Matched nasal epithelial samples from newborns were collected within six weeks from birth (15.9 ± 7.9 days [SD]) post-birth as detailed in the study protocol (1). All samples were cryopreserved at -80°C until DNA extraction was performed.

#### Nucleic Acid Isolation

Genomic DNA was extracted from both nasal brushings and amniotic membrane samples using the chemagic 360 automated nucleic acid isolation system (Revvity, Baesweiler, Germany) and the chemagic™ DNA Blood 400 Kit H96 (Revvity, part# CMG-1091), following the manufacturer's instructions, and then stored at -80 °C until analysis. DNA quantity was assessed using the Qubit HS dsDNA Assay (Q32854, Thermo Fisher Scientific) on a Qubit fluorometer (Thermo Fisher Scientific, Waltham, MA) (Figure E1 in the online supplement).

**DNA methylation measures**

Libraries were constructed using capture DNA methylation sequencing with enzymatic conversion (EM-seq) and target enrichment employing the TWIST Human Methylome Panel. Libraries were prepared from 200 ng of genomic DNA using the NEB Next Enzymatic Methyl-seq Library Preparation Protocol (Twist Bioscience and New England Biolabs, CA, USA) following the manufacturer's instructions. Targeted capture was performed using the Twist Targeted Methylation Sequencing protocol. Pre-capture libraries were pooled in 8-plex format and hybridized at 60°C for 16 hours. Subsequently, the hybridized pools were washed, and PCR amplified for six cycles according to the manufacturer's protocol. Finally, capture libraries were sequenced at the Genomics WA facility on an Illumina NovaSeq 6000 (Illumina, CA, USA) using a pair-end configuration with 150 base pair (bp) length reads (Figure E1 in the online supplement).

**Bioinformatic Analysis**

Bioinformatics analyses utilized computing and data resource provided by the Australian BioCommons Leadership Share (ABLeS) program (5) and Pawsey Supercomputing Research Centre (6, 7). Figure E1 summarizes the main steps carried out during the bioinformatics analyses. Briefly, raw methylation FASTQ files were processed using the nf-core/methylseq v2.3.0 pipeline. Methylseq pipeline was executed under Nextflow v23.04.2 (Di Tommaso et al., 2017) and the Human Genome Reference Consortium Human Build 38 (GRCh38) using the BWA-meth/MethylDackel workflow. Subsequent metrics and filtering were performed using target-methylseq-qc pipeline v2.1.0 (9), which included targeted methylation coverage assessment via *Picard-Profiler* mode and generation of targeted BED files based on BED intervals from the TWIST Human Methylome Panel using *bed-filter* mode. The Kids Research Institute Australia cluster and Google Cloud infrastructure were utilized for further analyses. Relevant scripts for data pre-processing are available on GitHub (https://github.com/wal-yan/AERIAL/tree/main/RespEpithelium_Methylome).

**Statistical Analyses**

Statistical analyses were conducted in R language v4.3.2 and RStudio v2023.03.0+386 (10). Figure E1 in the online supplement illustrates the flow of the analyses performed. Where specific packages are not mentioned, analyses were conducted using base R functions. During data quality control (QC), 2.58% of CpG sites (n=204,963) with zero coverage in any sample were removed. Additionally, 9.70% of CpG loci (n=770,067) with very low coverage (<5) or extremely high coverage (>500) were excluded. The remaining 87.72% of CpG sites (n=6,961,516) that passed QC were used for subsequent downstream analysis. Mitochondrial and non-standard chromosomes were removed from the dataset. Methylation ratios were derived from sequencing counts and expressed as beta (β) as follows:

$$\frac{methylated\ alleles}{(unmethylated + methylated) * 100}$$

With $\log_2$ transformation to *M*-values for statistical analysis. Sample quality control was performed by sex inference by extracting methylation calls on sex chromosomes and comparing to self-reported sex to identify potential sample mix-ups. Dimensional reduction was conducted using Principal Component Analysis (PCA) on M values (Figure E1, Step1, in the online supplement).

Differential methylation was evaluated both within regions (differentially methylated regions, DMRs) and at individual CpG sites (differentially methylated positions, DMPs) using linear regression. DMRs were detected using the DMRcate package v2.16.1 (11) and the model built using tissue type as the primary variable and adjusting only for gender, as DMRcate method corrects per sample using modelMatrixMeth function from edgeR package (12), therefore participant ID were not included to avoid overfitting. To define DMRs, a minimum of 50 CpG sites was used, applying a stringent threshold for genome-wide significance (Fold Discovery Rate (FDR) ≤ 0.01 and mean difference ≥ |1|) (Figure E1, Step2, in the online supplement). For CpG-level analysis, the limma package v3.58.1 (13) was used to construct a linear model with tissue type as the primary variable including gender as fixed effect. We also adjusted for the non-independence of samples from the same individual by applying the *duplicateCorrelation()* method from the limma v3.58 (13), using individual ID as a blocking variable. The estimated correlation was included during model fitting using *lmfit()* to improve the accuracy of differential analysis. Tissue-specific DMPs were identified using a threshold of FDR ≤ 0.05 and $\log_2$ Fold Change ($\log_2$FC) > |0.2| (Figure E1, Step3, in the online supplement). To identify conserved CpGs, we first retained loci with no significant differences between tissues (FDR ≥ 0.1). Among these, we then selected CpGs with high concordance in methylation levels between tissues (Pearson's correlation, R ≥ 0.8). Note that the FDR threshold refers to the differential methylation analysis between tissues, not the correlation with exposures. Gestational exposures (maternal smoking and maternal asthma) were then evaluated separately within conserved regions using the same limma-based linear modelling approach describe above, adjusting for sex and accounting for repeated measures by participant ID. Significant associations were defined as FDR ≤ 0.05 and $\log_2$FC ≥ |0.2| (Figure E1, Step 4, in the online supplement).

DMPs were annotated to genes using the annotatr v1.28.0 (14), or internal functions for DMR calling using DMRcate v2.16.1 (11). Gene Ontology (GO) pathway enrichment analysis was conducted using the rGREAT v2.4.0 (15), applying the 'Two nearest genes' rule where gene regulatory domains were defined by extending in both directions to the nearest transcriptional start site, but no more than 20kb in either direction. Statistically enriched pathway terms were summarized using the Simplify Enrichment package v1.12.0 (16). Supervised Partial-Least Square (PLS) Discriminant Analysis (DA) was implemented with mixOmics v6.26.0 (17) to assess associations with maternal asthma history and *in utero s*moking exposure. Visualization of BAM files was performed using the Integrative Genome Viewer (IGV) software v2.8.9 (18)

**Urinary Cotinine Levels**

Maternal smoking status was determined via positive response to questionnaire and validated with urinary cotinine levels (19) . Urinary cotinine levels were measured using the Salimetrics® High Sensitivity Salivary Cotinine Enzyme Immunoassay Kit (Cat No. 1-2002-5, SALIMETRICS, State College, Pennsylvania 16803, USA), following the manufacturer’s protocol. Urine samples collected at 20 and 36 weeks of pregnancy as part of the ORIGINS biobank protocol (2, 3) were preserved at -80°C until analyzed. A standard curve was then established using a 4-parameter non-linear regression curve fit, and cotinine concentrations calculated using GraphPad Prism software v9.3.1. The assay lower limit of cotinine detection was 0.15 ng/mL. Positive urinary cotinine tests were >10 ng/mL with those below 10 ng/ml as non-smokers.

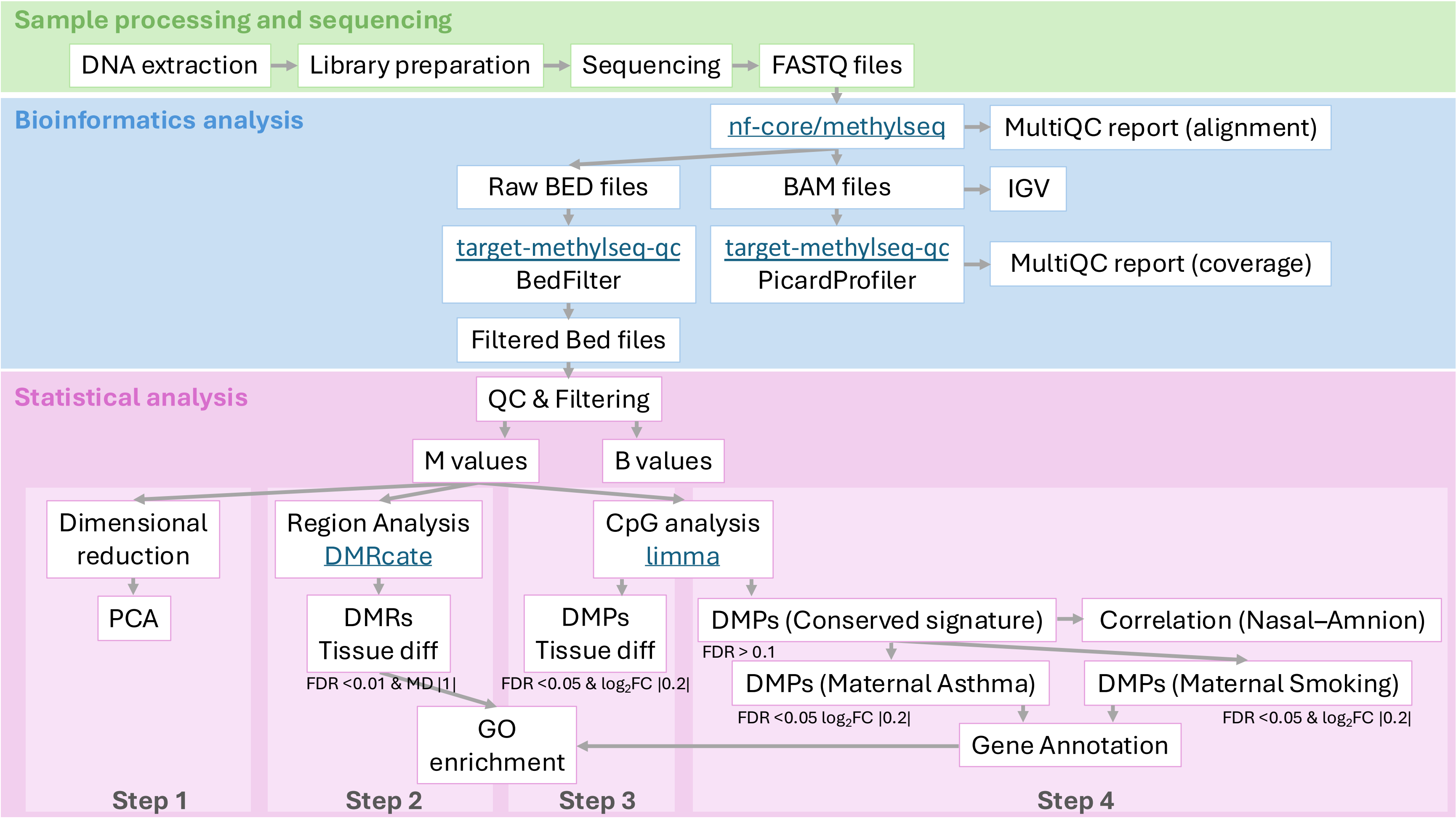


**Figure E1. Analysis Workflow.** This study was conducted in three main phases: Sample processing and sequencing (green), Bioinformatics analysis (blue), and Statistical analysis (pink). The statistical analysis phase was further divided into four key steps: Step 1 – Dimensional reduction using Principal Component Analysis (PCA); Step 2 – Differential methylation analysis at the regional level (differentially methylated regions, DMRs) associated with tissue differences; Step 3 – Differential methylation analysis at the CpG site level (differentially methylated positions, DMPs) associated with tissue differences; Step 4 – Identification of a conserved DMP signature, which was then used to assess associations with gestational exposures (maternal asthma and maternal smoking). MD: Mean Difference; FDR: False Discovery Rate; FC: Fold Change.

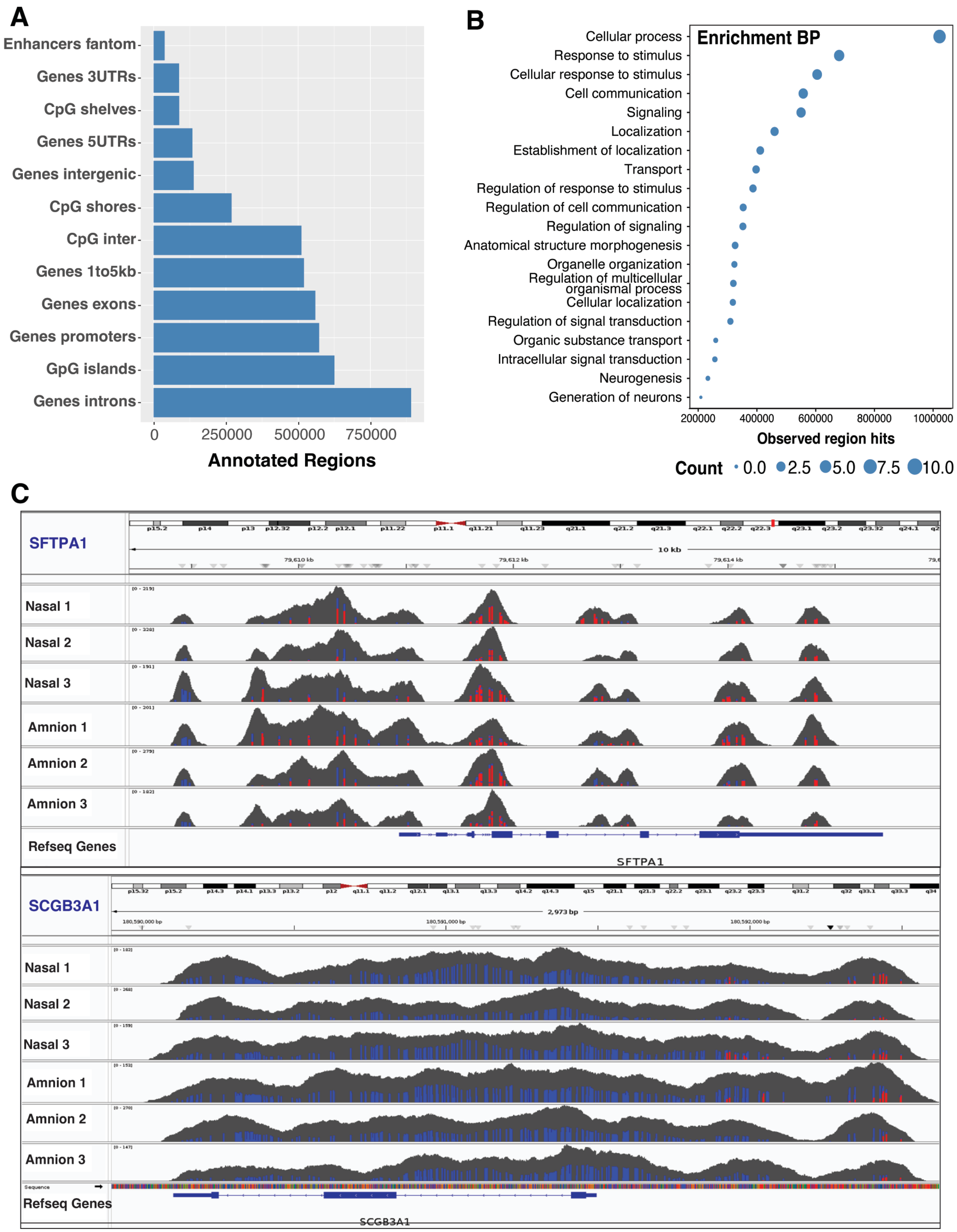
A
Enhancers fantom
Genes 3UTRs
CpG shelves
Genes 5UTRs
Genes intergenic
CpG shores
CpG inter
Genes 1to5kb
Genes exons
Genes promoters
GpG islands
Genes introns
0
250000
500000
750000
Annotated Regions
B
Enrichment BP
Cellular process
Response to stimulus
Cellular response to stimulus
Cell communication
Signaling
Localization
Establishment of localization
Transport
Regulation of response to stimulus
Regulation of cell communication
Regulation of signaling
Anatomical structure morphogenesis
Organelle organization
Regulation of multicellular organismal process
Cellular localization
Regulation of signal transduction
Organic substance transport
Intracellular signal transduction
Neurogenesis
Generation of neurons
200000
400000
600000
800000
1000000
Observed region hits
Count 0.0 2.5 5.0 7.5 10.0
C
SFTPA1
Nasal 1
Nasal 2
Nasal 3
Amnion 1
Amnion 2
Amnion 3
Refseq Genes
SFTPA1
SCGB3A1
Nasal 1
Nasal 2
Nasal 3
Amnion 1
Amnion 2
Amnion 3
Refseq Genes
SCGB3A1

**Figure E2. Conserved methylation landscape across tissues. (**A) Histogram counts of CpGs annotated to genomic features for the conserved methylome. (B) Significantly enriched biological processes within the conserved methylome for CpGs annotate to nearest transcriptional start sites. Point size represents count of number of genes relative to total genes within the pathway (C) Raw sequencing coverage across the *SFTPA1* gene coding region (top panel) and the *SCGB3A1* coding region. Ticks in red are methylated reads and blue represents unmethylated reads. Height of the grey silhouettes represents sequencing coverage.

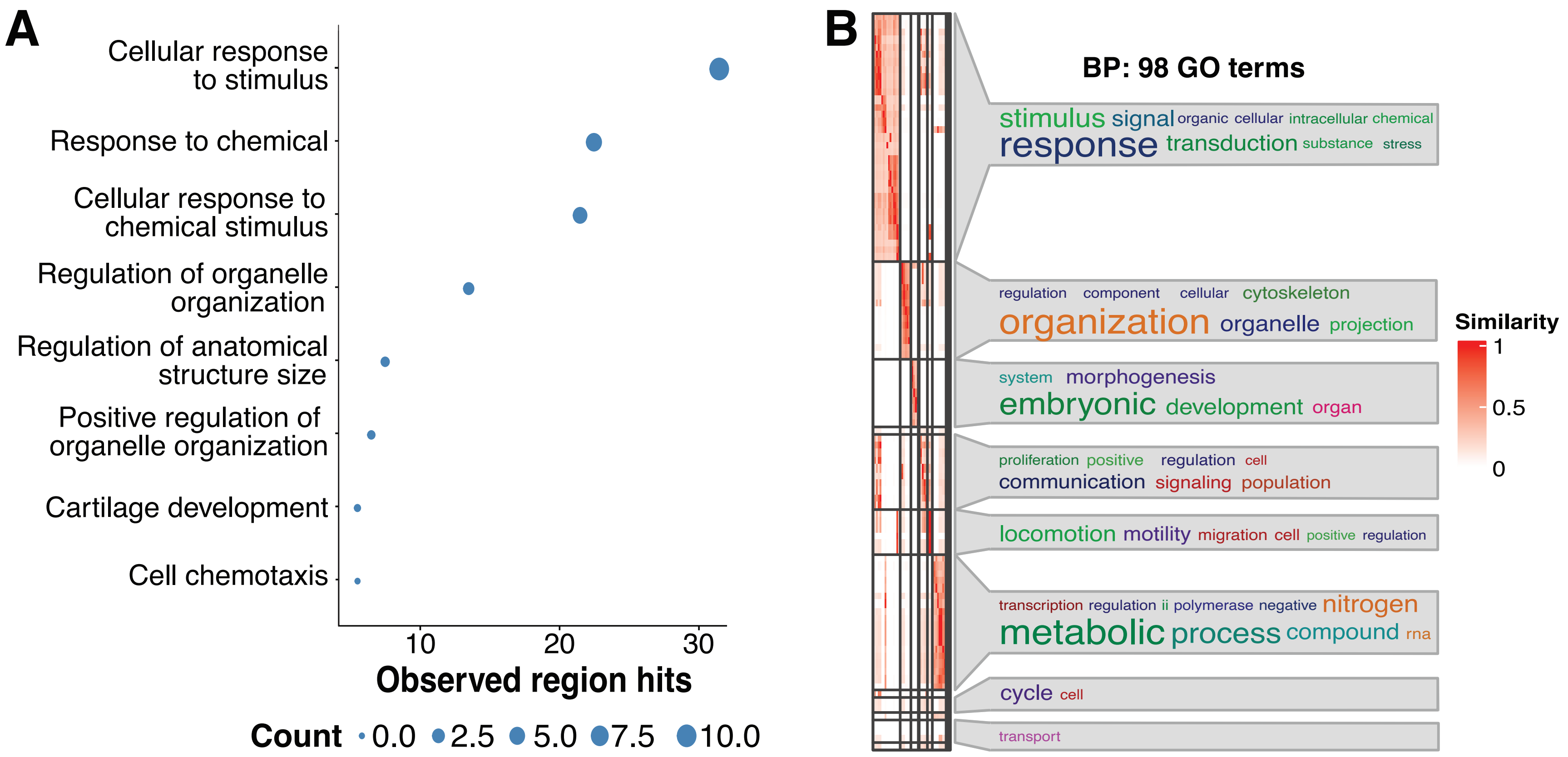


**Figure E3. Pathway enrichment of maternal asthma history.** (A) Significantly enriched biological processes within the conserved methylome for CpGs associated with maternal asthma. Point size represents count of number of genes relative to total genes within the pathway. (B) Gene Ontology (GO) enrichment was calculated using genes near CpG sites from the maternal asthma signature, followed by a similarity matrix analysis. The similarity heatmap displays GO terms associated with Biological Process (BP), with colour intensity representing the significance of enrichment.

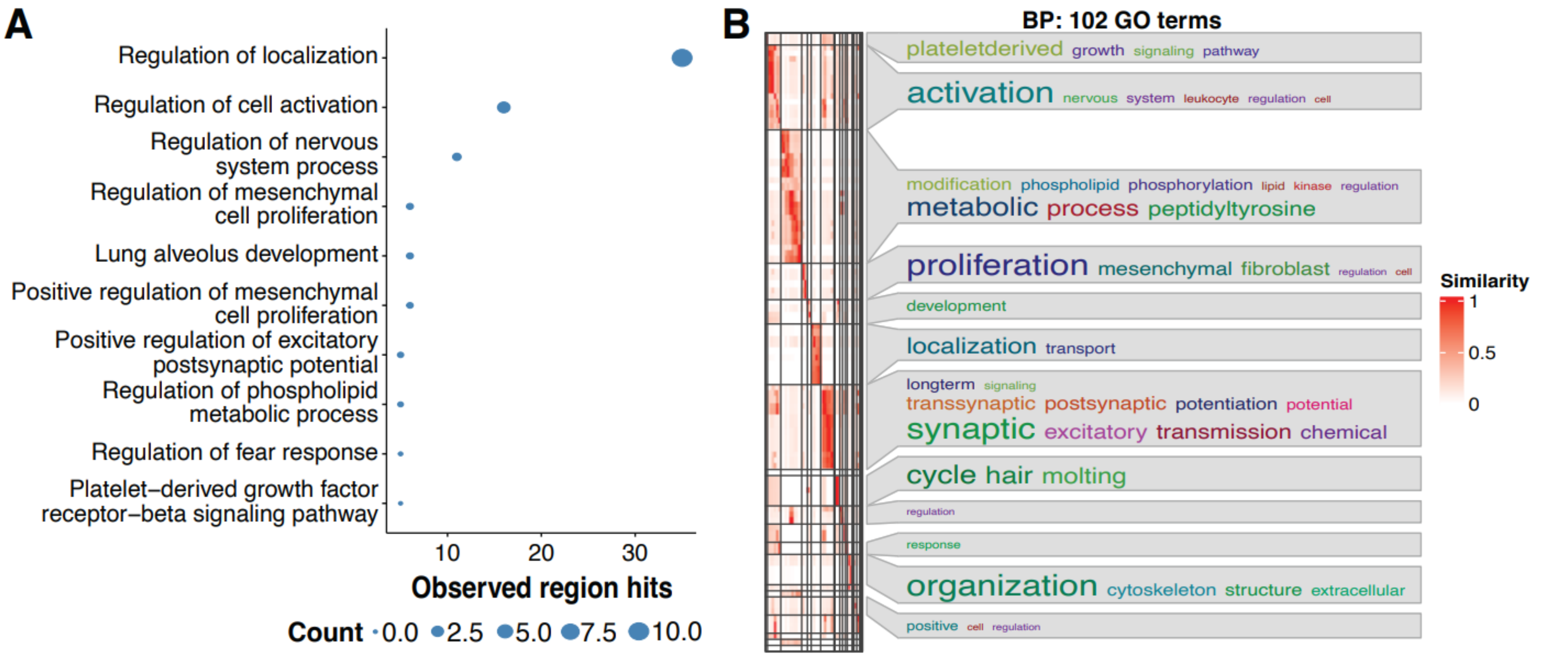


**Figure E4. Pathway enrichment of maternal smoking exposure.** (A) Significantly enriched biological processes within the conserved methylome for CpGs associated with maternal smoking. Point size represents count of number of genes relative to total genes within the pathway. (B) Gene Ontology (GO) enrichment was calculated using genes near CpG sites from the maternal smoking signature, followed by a similarity matrix analysis. The similarity heatmap displays GO terms associated with Biological Process (BP), with colour intensity representing the significance of enrichment.
